# LesionQuant for assessment of MRI in multiple sclerosis - a promising supplement to the visual scan inspection

**DOI:** 10.1101/2020.04.01.20048249

**Authors:** Synne Brune, Einar A. Høgestøl, Vanja Cengija, Pål Berg-Hansen, Piotr Sowa, Gro O. Nygaard, Hanne F. Harbo, Mona K. Beyer

## Abstract

**Background and goals:** Multiple sclerosis (MS) is a central nervous system inflammatory disease where magnetic resonance imaging (MRI) is an important tool for diagnosis and disease monitoring. Quantitative measurements of lesion volume, lesion count, distribution of lesions and brain atrophy have a potentially significant value for evaluating disease progression. We hypothesize that utilizing software designed for evaluating MRI data in MS will provide more accurate and detailed analyses compared to the visual neuro-radiological evaluation.

**Methods:** A group of 56 MS patients (mean age 35 years, 70% females and 96% relapsing-remitting MS) was examined with brain MRI one and five years after diagnosis. The T1 and FLAIR brain MRI sequences for all patients were analysed using the LesionQuant(LQ) software. These data were compared with data from structured visual evaluations of the MRI scans performed by a neuro-radiologist, including assessments of atrophy and lesion count. Correlations with clinical tests like the timed 25-foot walk test (T25FT) were performed to explore additional value of LQ analyses.

**Results:** Lesion count assessments by LQ and by the neuro-radiologist were significantly correlated one year (cor=0.92, p=2.2×10^−16^) and five years (cor=0.84, p=2.7×10^−16^) after diagnosis. LQ detected a reduction in whole brain percentile >10 in 10 patients across the time-points, whereas the neuro-radiologist assessment identified six of these. The neuro-radiologist additionally identified five patients with increased atrophy in the follow-up period, all of them displayed decreasing low whole brain percentiles (median 11, range 8-28) in the LQ analysis. Significant positive correlation was identified between lesion volume measured by LQ and test performance on the T25FT both at one year and five years after diagnosis.

**Conclusion:** For the number of MS lesions at both time-points, we demonstrated strong correlations between the assessments done by LQ and the neuro-radiologist. Lesion volume evaluated with LQ correlated with T25FT performance. LQ-analyses were more sensitive in capturing brain atrophy than the visual neuro-radiological evaluation. In conclusion, LQ seems like a promising supplement to the evaluation performed by neuro-radiologists, providing an automated tool for evaluating lesions and brain volume in MS patients in both a longitudinal and cross-sectional setting.

## 1 Introduction

Multiple sclerosis (MS) is a chronic inflammatory, demyelinating disorder of the central nervous system. Most frequently MS is characterized by a relapsing remitting disease course (RRMS) that over the years often converts to a secondary progressive disease course (SPMS). MS leads to variable degrees of physical and cognitive impairment. About 10 percent of the patients experience a progressive disease course from disease onset (primary progressive MS (PPMS)) (Koch et al., 2009;Lublin et al., 2014).

A key challenge in MS care is to identify and develop prognostic biomarkers for the disease course (Katsavos and Anagnostouli, 2013). Magnetic resonance imaging (MRI) is still the most important tool for the diagnosis and monitoring of MS (Wattjes et al., 2015;Filippi et al., 2016;Geraldes et al., 2018). MRI has a high sensitivity for the evaluation of inflammatory and neurodegenerative processes in the brain and spinal cord and it is the most commonly used method in the follow-up of MS patients (Kaunzner and Gauthier, 2017).

Visual inspection of the MRI scans of people with MS is time consuming for the neuro-radiologist. Subjective measurements based on a radiologist’s visual inspection may result in low reproducibility (Dwyer et al., 2017). In addition, the degrees of brain atrophy and MS-related pathology in the grey and white matter may be difficult to estimate, especially in early adulthood (Eshaghi et al., 2018;Storelli et al., 2018;Sastre-Garriga et al., 2020). New MRI post-processing tools that automatically analyse complex brain volumetric information and lesion load have recently become commercially available. A study comparing two different software types for assessment of longitudinal whole brain atrophy in MS patients found a strong level of statistical agreement and consistency between the two programs in a real-world MS population (Beadnall et al., 2019). The authors conclude that automated measurements of atrophy show promise as biomarkers of neuro-degeneration in clinical practice and will enable more rapid clinical translation. If these types of programmes are to be introduced, in addition to showing their performance compared to competing programs or established research tools, we need to evaluate their use compared to clinical practice today. Do they perform as well, or better than current practice, and in what way can they be useful and valuable?

Although a neuro-radiological evaluation of structural brain MRI in MS patients can easily estimate the pathologic burden of abnormalities such as T2 hyperintense lesions, limited correlation exists between these measures and the clinical phenotype (Mollison et al., 2017). This has been termed the ‘clinico-radiological paradox’ and is well described for both physical and cognitive impairments. Some explanations to this paradox have been suggested, including inappropriate clinical rating, and underestimation of damage to the normal appearing brain tissue (Barkhof, 2002). In a large meta-analysis including 2891 patients, Mollison et al found a modest correlation (*r=-* 0.30) between MRI measures of total brain white matter lesions and cognitive function in people with MS.

LesionQuant(LQ) by CorTechs Labs is a software that automatically segments and measures volumes of brain structures and compares these volumes to norms based on the more established NeuroQuant(NQ) software (Brewer, 2009). LQ was specifically designed for the evaluation of lesions and atrophy in MS patients. LQ also provides volumes and counts of new and enlarging brain lesions, and it automatically labels, visualizes and obtains the volumetric quantification of lesions based on brain MRI. LQ can therefore be used in the longitudinal follow-up of patients with MS. A recent study compared NQ to another software tool, Functional Magnetic Resonance Imaging of the Brain’s (FMRIB’s) Integrated Registration Segmentation Tool (FIRST), for estimating overall and regional brain volume in patients with clinically isolated syndrome (Pareto et al., 2019). To our knowledge, no data is published that compares the LQ software in MS with visual evaluations performed by neuro-radiologists.

By using data from our prospective longitudinal study of newly diagnosed MS patients, results from longitudinal LQ analyses were compared to visual evaluation performed by a neuro-radiologist in our hospital. We hypothesize that quantitative measurements of brain lesions and atrophy, using an unbiased automatic tool, may improve the correlation between clinical phenotype and MRI results.

Our aims were to evaluate a) The performance of LQ at detecting brain lesions compared to a neuro-radiologist, and b) brain atrophy as measured by LQ in comparison to the visual inspection by the neuro-radiologist. c) the correlations of results from both visual assessment and LQ with clinically relevant variables and d) the correlation between the segmented brain volumes acquired from LQ with the established FreeSurfer method.

## 2 Materials and methods

### 2.1 Participants

All analyses were based upon carefully phenotyped MS patients in an ongoing prospective longitudinal MS study in Oslo (Nygaard et al., 2015a;Høgestøl et al., 2019). A total of 56 MS patients were included in this study, which had been examined on average one year after diagnosis. The inclusion criteria were a diagnosis of RRMS in the period 2009-2012 and age 18-50 years. The exclusion criteria were a history of psychiatric or other neurological diseases than MS, drug abuse, previous adverse Gadolinium reaction, pregnancy or breast-feeding at inclusion, or non-fluency in Norwegian.

Data from two time-points after diagnosis of MS were included in this study; data from time-point 1 (TP1) was collected 13 months after diagnosis (±9, n=56) and data from time-point 2 (TP2) after 66 months (±12, n=56). At both time-points an expanded disability status scale (EDSS) score was calculated based on a standard neurological examination by trained clinicians (Kurtzke, 1983;Cutter et al., 1999). For assessment of walking ability and upper extremity function we also performed timed 25-foot walk test (T25FT) and 9-Hole Peg Test (9HPT). A brain MRI scan for clinical and research setting, was performed at both time-points.

We classified the disease modifying treatments (DMTs) as follows; group 0: no treatment, group 1: Glatiramer Acetate, Interferons, Teriflunomide or Dimetylfumarate, group 2: Fingolimod, Natalizumab or Alemtuzumab.

### 2.2 MRI acquisition

MS patients were scanned at both time-points with the same MRI scanning protocol in the same 1,5 T scanner (Avanto, Siemens Medical Solutions; Erlangen, Germany). The following two MRI sequences were required for the LQ analyses in this study;

a. A Sagittal 3D T1 MPRAGE (FOV: 240 × 240 mm; slice thickness: 1.2 mm; voxel size: 1.3 × 1.3 × 1.2 mm; TR: 2400 ms; TE: 3.61 ms; TI: 1000 ms; flip angle: 8 deg.
b. Pre-contrast sagittal 3D FLAIR (FOV: 260 × 260 mm; slice thickness: 1 mm; voxel size: 1 × 1 × 1 mm; TR: 6000 ms; TE: 333 ms; TI: 2200 ms.

The neuro-radiologist could in addition use all available other sequences in the study protocol, as mentioned in previous publications (Nygaard et al., 2015a;Nygaard et al., 2015b).

### 2.3 LesionQuant analyses of brain volume and lesion count

The MRI data from the 56 patients were analysed using the LQ tool (version 2.3.0), comparing data at TP1 and TP2. For each patient, T1-weighted and FLAIR sequences were uploaded to the LQ server from the PACS system in the hospital, without the need for pre-processing. Finalized LQ-reports were received after approximately 10 minutes. The reports provided volumes and counts of all lesions, including new and enlarging lesions at TP2. Volumes of brain structures in the MS patients were compared with age and sex matched healthy controls from the LQ reference database. To establish the normative LQ database, CorTechs Labs combined data from several thousand scans including publicly available studies, studies by collaboration partners, and other proprietary data sources (age range 3-100 years, acquired using Siemens, GE and Philips MRI scanners with both 1.5T and 3T field strength).

We performed a manual lesion count of all lesions at TP1 and TP2 to compare with the exact number of lesions counted by LQ. The LQ-reports provided information about volumes of different brain structures, including whole brain, thalamus, cerebral white matter, volume of white matter lesions and cortical grey matter. The results for each patient are illustrated both using percentiles and absolute values (Fig. 1). A cut off for atrophy was defined as a 10 percentile drop in brain volume for LQ between TP1 and TP2 (= five-year interval). This was set higher than expected reduction in brain volume per year as published in previous papers (Battaglini et al., 2019;Sastre-Garriga et al., 2020) to increase sensitivity, since we compared the results to a visual evaluation. Advanced tools have been shown necessary to detect early brain atrophy in MS (Filippi, 2015). To compare LQ with a well-known and validated research method for brain segmentation we used the measure for whole brain volume (excluding brain stem) from both LQ and FreeSurfer (Dale et al., 1999).

**Figure 1.**
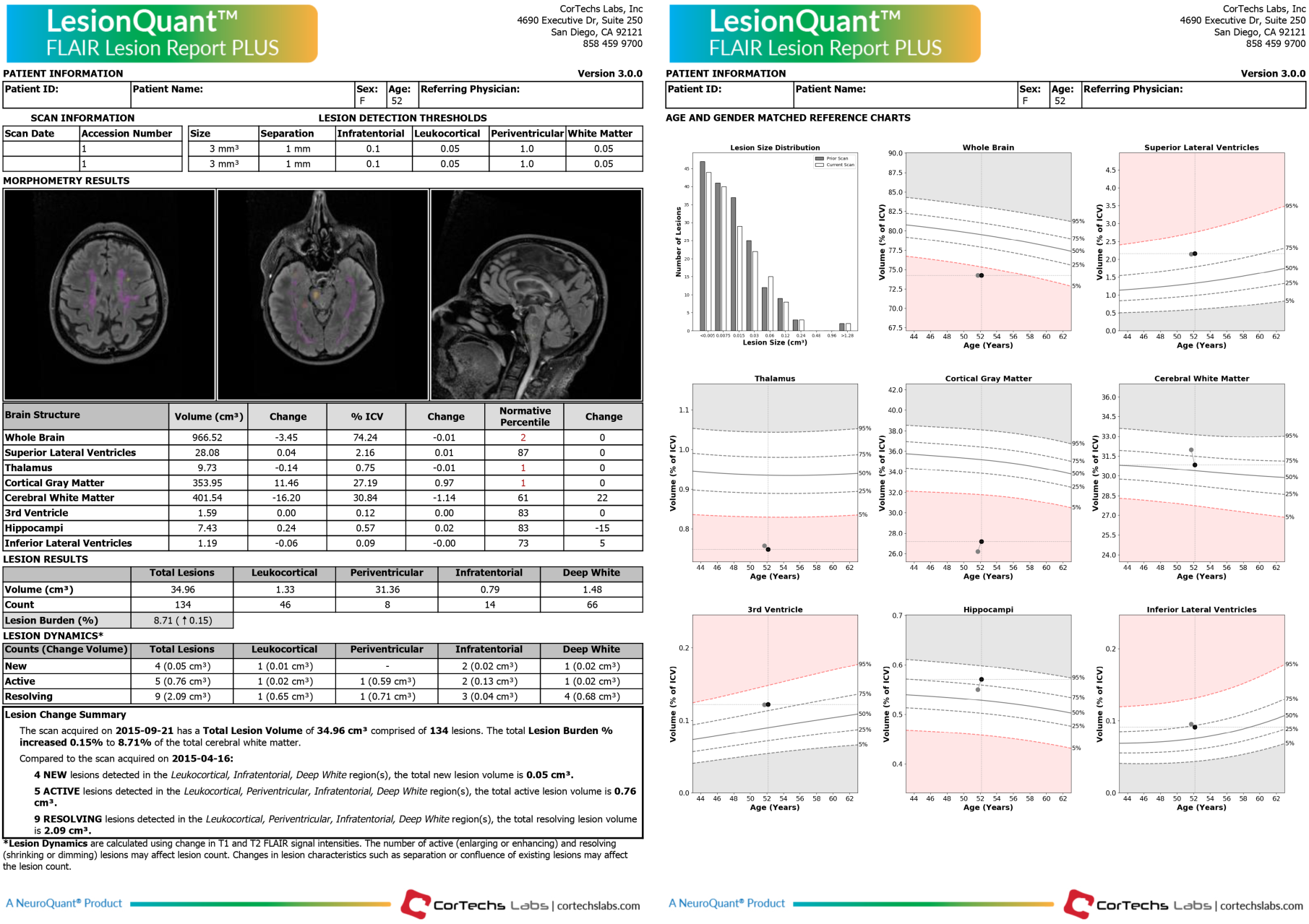
LesionQuant report. Example of a LesionQuant report from one MS subject comparing two MRI scans with a five-month time interval between the two time-points.

### 2.4 MRI evaluation of brain volume and lesion count presented by the neuro-radiologist

The brain MRIs from the included MS patients were systematically evaluated by a neuro-radiologist who carefully counted all lesions on the MRI scans of the patients at TP1 and TP2, and assessed whether increased whole brain atrophy was visually evident between the two time-points (Figure 2). Visual evaluation of atrophy was done using the 3D T1 series, where increased CSF in the sulci or on the surface of the brain or volume loss of the gyri between TP1 and TP2, was regarded as atrophy. A lesion was defined as having a high T2/FLAIR signal ≥ 3 millimetres in diameter.

**Figure 2.**
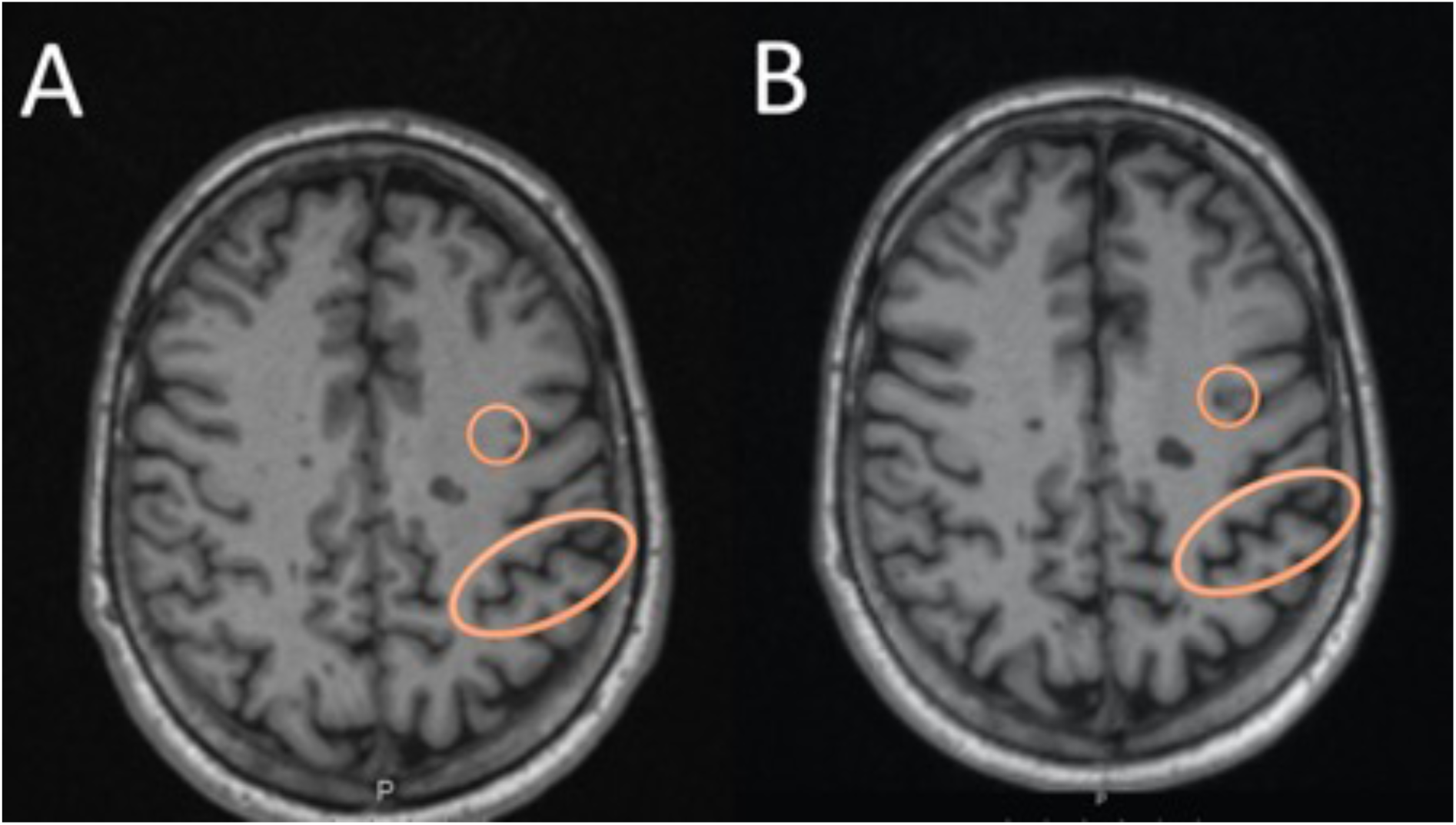
An example of the visual assessment by the neuro-radiologist. In (A) we see an axial T1 MRI at time-point 1, while in (B) we see the MRI at time-point 2, highlighting a circle with an example of a new lesion evolving during the follow-up period. The oval circle is an example of an area showing increased CSF in the sulcus, which was evaluated as representing atrophy between the two time-points.

**Figure 3.**
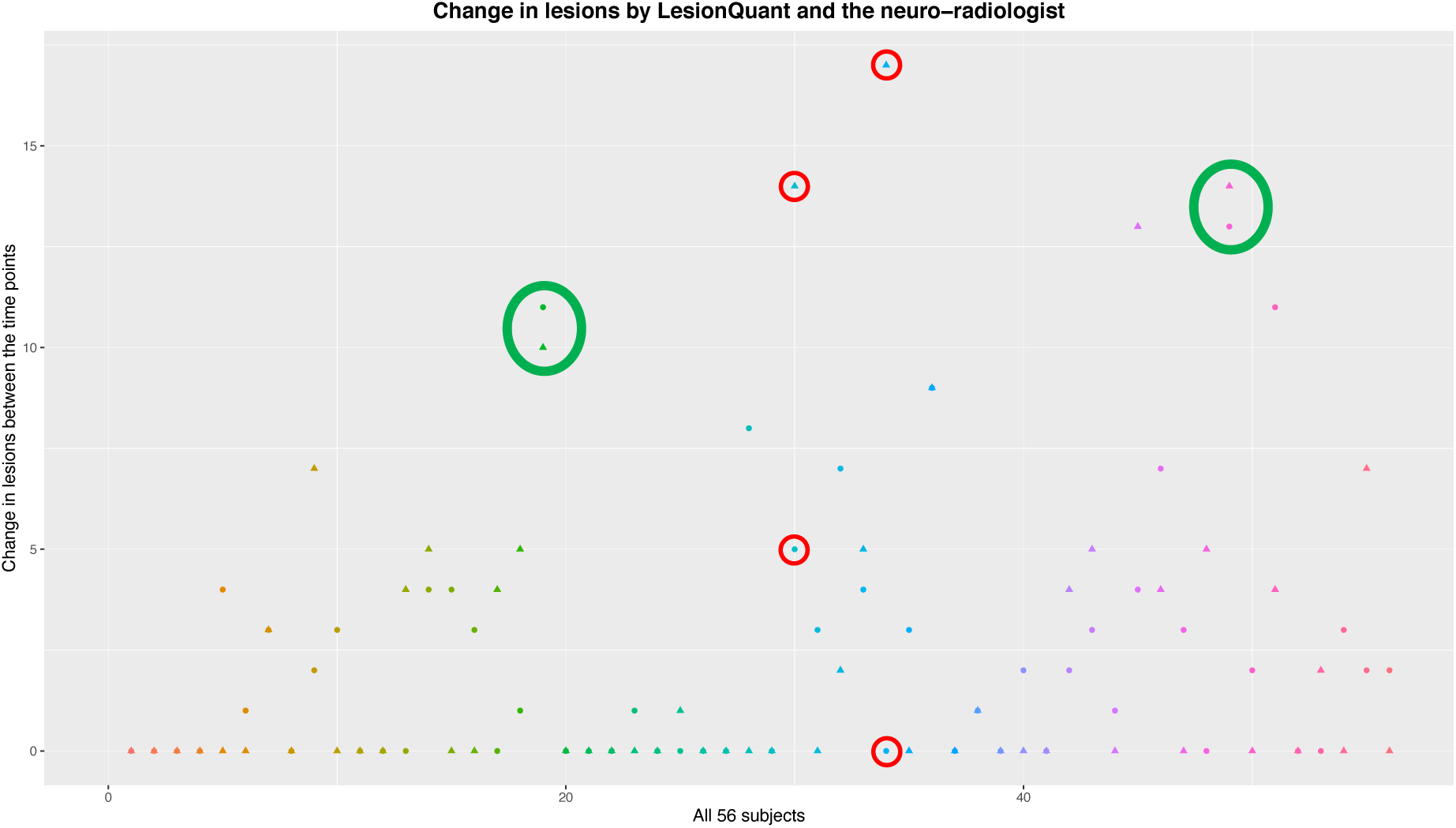
An overview of the evaluations of change in lesions between the two time-points. The LesionQuant assessments are depicted with a circle, while the neuro-radiological evaluations are depicted using a triangle. Each subject is visualized with both assessments and with a unique colour. The green circles show examples of assessments with good agreement between LQ and neuro-radiologist, while the red circles show assessments where the two methods differ a lot in the same patient.

The exact lesion number, and evaluation of atrophy was then compared to the output from LQ. The assessment by the neuro-radiologist was used as the gold standard to compare the LQ data with.

### 2.5 MRI pre and post-processing using FreeSurfer

The MRI pre and post-processing for FreeSurfer analyses of these data has been described previously (Høgestøl et al., 2019). MRI variables were acquired from the FreeSurfer output stats. We used the variable “Brain Segmentation Volume Without Ventricles from Surf” as measure of total brain volume, which excludes the brainstem.

### 2.6 Statistical analysis

We used R (R Core Team, Vienna, 2018, version 3.6.1) for statistical analyses. To assess reliability of the whole brain volumes from LQ and FreeSurfer we computed the intraclass correlation coefficient (ICC) using the R package “irr” (Gamer, 2019). Figures were made using “ggplot2” (Wickham, 2016) and “cowplot” (Wilke, 2019) in R.

To evaluate the associations between the assessment provided by LQ, the neuro-radiologist, analysis using the FreeSurfer software and the clinical data, we used the “stats” package in R and calculated the Pearson’s correlation coefficient and applied the student’s t-test (R Core Team, 2019).

To adjust for multiple comparisons, we calculated the degree of independence between the four clinical variables available, making a 4 × 4 correlation matrix based on the Pearson’s correlation between all pair-wise combinations of clinical data. Utilizing the ratio of observed eigenvalue variance to its theoretical maximum, the estimated equivalent number of independent traits in our analyses was 3.0 (Li and Ji, 2005). To control for multiple testing, our significance threshold was therefore adjusted accordingly from 0.05 to 0.017 (Li and Ji, 2005).

## 3 Results

### 3.1 Participant demographics and characteristics

At TP1 mean age of the study participants was 36 years (range 21-49 years), 70% were females and 96% were classified as having RRMS. EDSS was stable between TP1 and TP2 with median EDSS 2.0 (range 0-6). Time since MS diagnosis was on average 12.9 months (SD=9.3) at TP1 and 66.0 months (SD=11.7) at TP2. At TP1, 25% did not receive any DMT for MS, 63% received a group 1 DMT (moderately effective treatment) and 12% a group 2 DMT (highly effective treatment). At TP2, 34% did not receive any DMT, 36% received a group 1 DMT and 30% a group 2 DMT (Table 1).

**Table 1.**
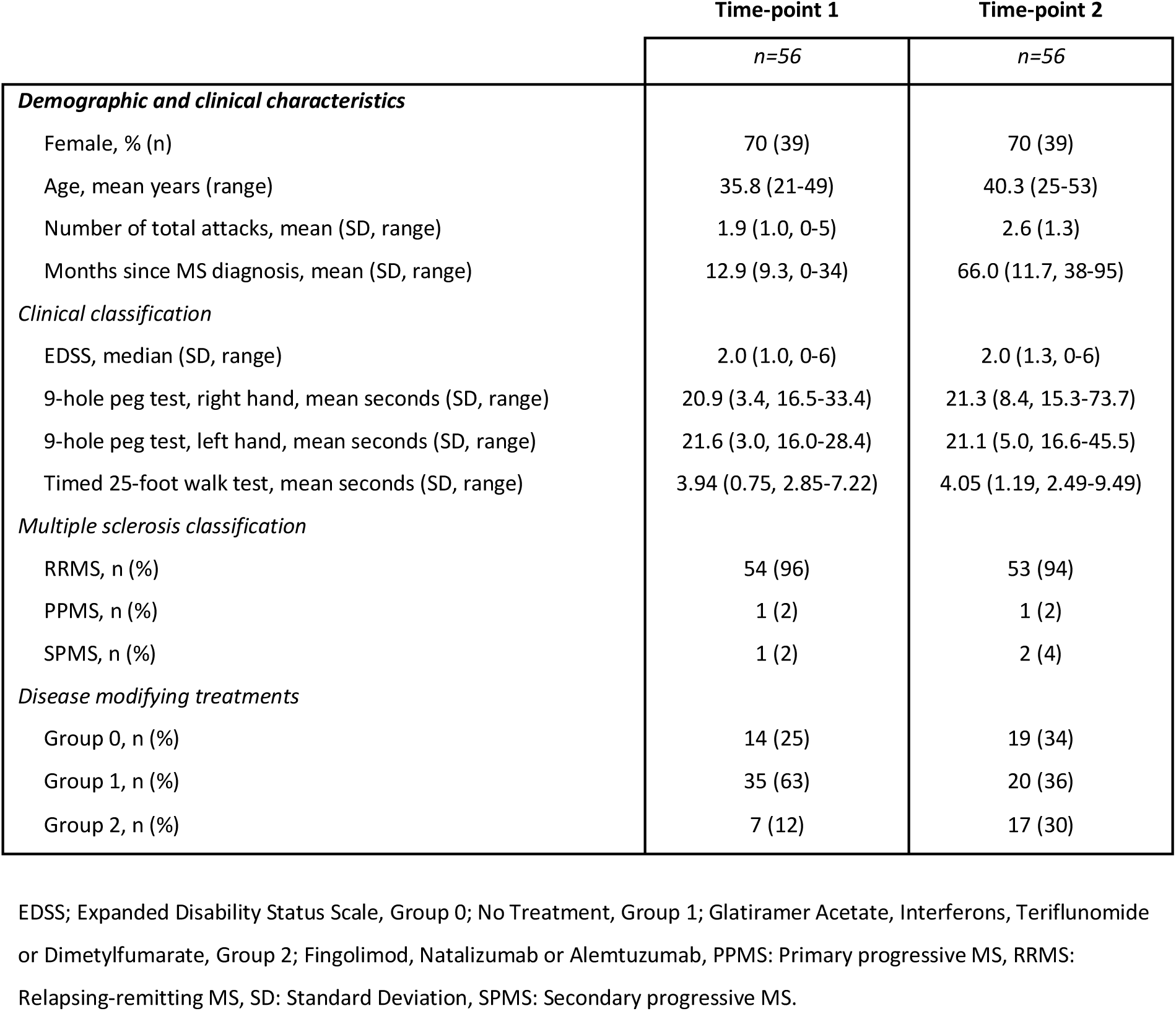
Demographic and clinical characteristics of the multiple sclerosis patients.

### 3.2 Comparison between the LesionQuant reports and the neuro-radiological evaluations

The lesion count assessments by LQ and the neuro-radiologist were significantly correlated at TP1 (cor=0.92, p=2.2×10^−16^) and TP2 (cor=0.84, p=2.7×10^−16^) (Fig. 2, Supplementary Fig. 2). The lesions counts were identical in only two and three patients at TP1 and TP2, respectively. While lesion counts were higher by LQ in 39 and 40 patients at TP1 and TP2, respectively.

Lesion counts were lower by LQ in 15 and 13 patients at TP1 and TP2, respectively. In general, the differences in number of lesions evaluated by LQ and the neuro-radiologist increased with age. For patients with lower number of lesions the neuro-radiologist tended to count more lesions than LQ, and the opposite with higher number of lesions, see Supplementary figure 1.

### 3.3 Longitudinal evaluations of lesions and atrophy

We also compared the LQ software with the assessment by the neuro-radiologist in identifying whole brain atrophy at TP2. The neuro-radiologist classified 12 subjects to have brain atrophy. These 12 subjects also had significantly lower scores on whole brain atrophy by LQ (mean brain volume percentile 37.0, range 10-80), compared to the subjects that were not classified as having brain atrophy (mean brain volume percentile 48.7, range 2-99).

At TP2, LQ and the neuro-radiologist agreed in classifying 33% of the subjects with atrophy (four out of 12 subjects). In addition, the neuro-radiologist identified eight more subjects with brain atrophy (mean LQ whole brain percentile 31.3).

LQ detected a reduction in whole brain percentile >10 in 10 patients between TP1 and TP2, while the neuro-radiological evaluation identified six of these. The evaluation by the neuro-radiologist identified an additional six patients with increased atrophy between TP1 and TP2, all of whom displayed low whole brain percentiles at TP2 (median 11, range 8-28) and decreasing percentile between the time-points.

At TP2 we found that LQ showed reduced whole brain volume in 51 patients compared to TP1 with a mean reduction in volume of 20.5 ml / 1.59% (range 0.4-109.4 ml / 0.03-8.08%) of the whole brain volume. In the remaining five patients we found an increased volume with a mean increase in volume of 6.8 ml/ 0.56% (range 0.2-17.4 ml / 0.02-1.44%).

To evaluate the sensitivity of LQ in detecting new lesions, compared to the neuro-radiologist, the difference in number of lesions assessed at the two time-points was analysed in a 2×2 table (Table 2). The sensitivity of the LQ-analysis to correctly classify the patients according to the gold standard neuro-radiological evaluation was 53% (17/32 patients). The specificity of the LQ-analysis to correctly evaluate the MRI follow-up as stable according to the neuro-radiological evaluation was 75% (18/24). In total, 43 % of the patients were evaluated with no new lesions on MRI at TP2 by the neuro-radiologist. Also, 57% (32 patients) had new lesions according to the neuro-radiologist, and only 17 of these had new lesions according to the LQ-reports (Table 2, Fig. 2).

**Table 2.** A 2×2 table based on the ability to capture MRI activity based on the presence of new lesions in our longitudinal MS sample.

|  |  | LesionQuant report |  | In total |
| --- | --- | --- | --- | --- |
|  |  | ***New lesions*** | ***Stable*** |  |
| Neuro-radiological evaluation | ***New lesions*** | 17 | 15 | 32 |
|  | ***Stable*** | 6 | 18 | 24 |
|  | ***In total*** | 23 | 33 | 56 |

### 3.4 Correlations between MRI features and clinical variables

We found significant positive correlations between T25FT and the lesion volume as measured by LQ at both TP1 (t=3.08, p=3.2 × 10^−3^) and TP2 (t=3.72, p=4.8 × 10^−4^) (Table 3). These results also indicate slower test performance by T25FT in patients with a higher burden of lesion volume. In addition, we found a significant positive correlation between the 9HPT using the left hand and lesion volume at TP2 (t=5.34, p=2.09 × 10^−6^), indicating slower test performance with increased lesion volume. We also found a significant negative correlation with EDSS and whole brain volume at TP1 (t=-2.68, p=9.8 × 10^−3^), indicating higher EDSS scores with lower brain volumes. We found no significant correlations between the number of lesions reported by the neuro-radiologist and the clinical variables. All significant correlations reported were adjusted for multiple testing.

**Table 3.** Associations between LQ-variables and clinical variables. Results marked in bold and italics were significant after adjusting for multiple testing.

| Clinical variables | LesionQuant variables |  |  |  |
| --- | --- | --- | --- | --- |
|  | Lesion volume |  |  |  |
|  | Time-point 1 |  | Time-point 2 |  |
|  | t-value | p-value | t-value | p-value |
| Expanded disability status scale | 1.12 | 0.27 | 1.43 | 0.16 |
| 9-hole peg test, right hand | 2.00 | 0.05 | 1.87 | 0.07 |
| 9-hole peg test, left hand | 1.00 | 0.32 | **5.34** | ***2.09 x 10<sup>-6</sup>*** |
| Timed 25-foot walk test | **3.08** | ***3.2 x 10<sup>-3</sup>*** | **3.72** | ***4.8 x 10<sup>-4</sup>*** |
|  | Lesion count |  |  |  |
| Expanded disability status scale | 0.85 | 0.40 | 0.42 | 0.68 |
| 9-hole peg test, right hand | 2.35 | 0.02 | 0.50 | 0.62 |
| 9-hole peg test, left hand | -0.23 | 0.82 | 1.90 | 0.06 |
| Timed 25-foot walk test | 1.48 | 0.15 | 0.68 | 0.50 |
|  | Whole brain volume |  |  |  |
| Expanded disability status scale | **-2.68** | ***9.8 x 10<sup>-3</sup>*** | -1.45 | 0.15 |
| 9-hole peg test, right hand | 0.94 | 0.35 | 0.86 | 0.40 |
| 9-hole peg test, left hand | -2.24 | 0.03 | -0.20 | 0.84 |
| Timed 25-foot walk test | -0.68 | 0.50 | -1.22 | 0.23 |
|  | Whole brain percentile |  |  |  |
| Expanded disability status scale | -0.11 | 0.91 | -0.81 | 0.42 |
| 9-hole peg test, right hand | 1.05 | 0.30 | -0.33 | 0.74 |
| 9-hole peg test, left hand | -1.16 | 0.25 | -0.69 | 0.49 |
| Timed 25-foot walk test | 1.06 | 0.29 | 0.18 | 0.86 |
|  | Lesion count by neuro-radiologist |  |  |  |
| Expanded disability status scale | 1.00 | 0.33 | 1.06 | 0.30 |
| 9-hole peg test, right hand | 2.64 | 0.01 | 0.76 | 0.45 |
| 9-hole peg test, left hand | 0.05 | 0.96 | 2.69 | 0.01 |
| Timed 25-foot walk test | 1.11 | 0.27 | 1.20 | 0.24 |

### 3.5 Reliability of LesionQuant volumes

To validate the LQ data with the established FreeSurfer output for brain segmentation we compared the measure for whole brain volume (excluding brain stem) from both LQ and FreeSurfer, (Supplementary Fig 3). At both TP1 (t=51.6, cor=0.99) and TP2 (t=45.2, cor=0.99), Pearson’s correlations were highly significant. We also validated regional volumes for thalamus using both raw FreeSurfer data and data processed through the longitudinal stream compared to the LQ data, yielding less significant correlations (t=2.4-2.8 and, cor=0.32-0.35, p=0.02-0.008).

### 3.6 LesionQuant reports and neuro-radiological evaluation

All longitudinal LQ-data yielded excellent concurrence. To evaluate the consistency and agreement of the longitudinal LQ-reports, we measured the Intraclass correlation coefficients (ICC) between TP1 and TP2 for brain volume (ICC=0.97, p=2 × 10^−35^), lesion count (ICC=0.91, p=1.4 × 10^−23^), lesion volume (ICC=0.88, p=5.2 × 10^−20^) and thalamus volume (ICC=0.91, p=3.0 × 10^−23^) (Supplementary Table 1 and Supplementary Fig. 4). We found significant correlations between lesion volume and the number of lesions at both TP1 (t=6.32, p=5.05 × 10^−8^) and TP2 (t=4.21, p=9.77 × 10^−5^). We found no significant changes in the parameters between TP1 and TP2 (Supplementary Table 1). As a sanity check, the ICC for lesion counts reported by the neuro-radiologist was very high (ICC=0.99, p=2.6 × 10^−50^), as expected.

## 4 Discussion

Magnetic resonance imaging is an important para-clinical tool for the diagnosis and monitoring of MS. Quantitative measurements of lesion volume and distribution of lesions have a significant value for evaluating disease progression in a clinical setting and brain atrophy is a possible new measurement to be used in future evaluation in MS patients. In this study we explored the use of the LQ software for evaluating cerebral MS lesions as well as brain atrophy in a clinical setting, and investigated if an automatic analysis of MRI scans using such software shows promise for use in the clinical follow-up of MS-patients.

We found a high correlation between lesions counted by the neuro-radiologist at TP1 and TP2 and the lesion count output from LQ. In evaluation of atrophy between TP1 and TP2 there was only agreement between the neuro-radiologist and LQ in 50% of the patients (6 out twelve). Differences in whole brain percentiles between TP1 to TP2 were detected with LQ in the majority of patients, ranging between 0.03-8.08 %. Lesion volume from LQ analysis correlated with outcome of clinical tests of walking speed and upper extremity function. Significant positive correlation was identified between lesion volume measured by LQ and test performance on the T25FT both at one year (t=3.08, p=3.2 × 10^−3^) and five years after diagnosis (t=3.72, p=4.8 × 10^−4^).There was a significant correlation between the results of LQ and the segmented volumes by FreeSurfer, showing high reliability of LQ output for whole brain volume.

In order to evaluate treatment-effect, it is of importance to see if new or enlarging lesions appear on a follow-up MRI scan. The lesion counts of the LQ software and the neuro-radiologist were highly correlated at both timepoints. However, LQ assessment revealed somewhat higher lesion counts than the visual assessment, more so in patients with a high number of lesions. This could indicate a higher sensitivity to identify demyelinating lesions by the LQ software when there are many lesions. Many lesions are a challenge to the neuro-radiologist, as it is more difficult to keep track of lesion count. Another possible explanation is that unspecific lesions, wrongfully labelled as MS lesions are counted by LQ. With the high correlation of lesion count overall, the LQ tool should be valuable for detecting lesions in routine follow-up MRI in MS. The resulting LQ report could then be controlled by a neuro-radiologist to ensure that only probable MS lesions are included.

Regarding the detection of lesions, we used the assessment by the neuro-radiologist as the “gold standard”. However, it is well established that the detection of cortical lesions may be challenging using conventional brain MRI and these may be missed by radiologists (Patel et al., 2015). This is shown in a study comparing the number of MS lesions counted by radiologists and as analysed by MSmetrix (today known as IcoBrain MS), a software comparable to LQ (Jain et al., 2015). This study showed a higher recount-difference when recounting was performed by radiologists than in MSmetrix (D. Sima and Lysandropoulos, 2018). Therefore, the gold standard as we defined it in this paper, may be more variable than the automated software tool.

Reliable evaluation of atrophy is difficult with only visual inspection, although it is not a very time-consuming task. When the neuro-radiologist evaluated the MRI, images from TP2 was compared with the MRI scan at TP1 for each patient. In a clinical routine setting, the neuro-radiologists often compare to the previous MRI, which may be taken months or up to a year before. The changes in atrophy are rather small from year to year (-0.2 % to -0.3 % per year in our patients’ age range) (Battaglini et al., 2019) and it is not possible to detect such small changes in reduction of brain volume for the neuro-radiologist. A better approach may be to always compare the last scan to the first MRI in order to increase sensitivity of visual atrophy assessment. Results from studies comparing visual ratings of atrophy using GCA have shown Inter-rater reliability of > 0.6 and Intra-rater reliability of >0.7, which is considered moderate agreement (Pasquier et al., 1996). The discrepancy between the 12 patients found to have atrophy from visual inspection, to the 51 patients showing reduced brain volumes in the 5-year follow-up indicates that LQ would be helpful in clinical practice.

Most of the MS patients were treated with moderately or highly efficacious disease modifying therapies at TP1 and TP2. In total, 10 MS patients changed to a more efficient MS treatment during the follow-up. We found no significant differences in brain volumes or change in brain volumes between the patients who increased treatment efficiency during the follow-up and those who either used the same treatment or reduced the efficacy of their MS treatment during the follow-up period. As a confounding factor we have to consider that switching to more efficacious treatments would impact the brain volumes by possible pseudoatrophy during the first six months (De Stefano and Arnold, 2015). Although, during our observational period we did not find any significant differences. Other short term confounding factors affecting brain volume measurements are known, such as fluid restriction, the time of the day for MRI measurements, corticosteroids, antipsychotic treatment and short-term effects of pathological processes that decrease global brain volume.(Dieleman et al., 2017).

LQ compared differences in brain volume during approximately a five-year period (2012/2013 and 2016/2017). During this period the patients were scanned on several occasions, which were not part of the study. One of the main benefits of using automated methods for image analysis in MS patients, is the possibility to perform more reliable and quick evaluation of brain atrophy. As shown by Pareto et al, when comparing two different tools for automated volume analysis of different brain regions, the size of the brain region of interest seems to be important (Pareto et al., 2019). We found an excellent correlation between the FreeSurfer and LQ software’s in the assessment of whole brain atrophy (cor=0.99). However, for thalamus we only found modest correlations between both the raw and processed volumes estimated by LQ and FreeSurfer (cor=0.32-0.35, p=0.02-0.008, respectively), confirming the results of Pareto et al. In a recent paper, Storelli et al. also studied reproducibility and repeatability using different software’s for atrophy measurements (Storelli et al., 2018). They concluded that an improved reproducibility between scanners is required for clinical application.

In our study the LQ software estimated an unexpected increased whole brain volume in six patients between TP1 and TP2. This could be due to variability in the MRI scanner or other technical reasons. Alternatively, changes in lesion load in the patient over time may affect the atrophy measurements (Storelli et al., 2018). Overall, our results indicate that the automated method LQ performs better than the visual evaluation method in terms of atrophy evaluations, as discussed above.

We hypothesized that improved measurements of brain lesions and atrophy, using an unbiased automatic tool, may improve the correlation between clinical phenotype and MRI results. We found that only the automated LQ software was able to show significant correlation with the standard clinical tests (T25FT, 9HPT and EDSS). We consider this to be a robust and expected finding as only LQ and not the neuro-radiologist could provide volumetric information. In line with this, the 9HPT was positively correlated with lesion volume at time-point two; although only significant for the left hand, the same trend was seen for the right hand. The EDSS scale, which is the most widely used method to grade disability in MS, was only associated with whole brain volume at TP1 (t=-2.68, p=9.8 × 10^−3^). There were no correlations between lesion count, either from LQ or the neuro-radiologist and the EDSS, T25FT or 9HPT, also showing the value of having volumes of the lesions and whole brain available.

In general, we found very high levels of intraclass correlation coefficients (0.88-0.97), showing consistency and agreement among the longitudinal LQ-reports. A strength of this paper is the longitudinal design where the MS patients were examined clinically and with brain MRI both one and five years after diagnosis. The patient cohort is well characterised by trained clinicians, performing the clinical and MRI assessments. The same MRI scanner and protocol was used for all patients at the two time-points of evaluation, and all patients were scanned in the afternoon/early evening. The neuro-radiological evaluation at TP1 and TP2 was performed by a single neuro-radiologist. Thus, the quality of the data included in this study is suitable for addressing the research question. A weakness of the study is that we did not perform visual assessment by two independent raters for the visual evaluation. Also, there was no control group.

The structured LQ report is acquired using fully automated MRI post-processing software, which requires only minimal effort and reduces bias of different raters, which is present when using visual inspection of images. Another advantage is the very short processing time of LQ compared to similar software used for research, with only about 10 minutes from the uploading of images to the final report is received. In comparison, software like FreeSurfer needs hours to process the data, cannot be interpreted for individual patients and is not feasible for clinical practice.

## 5 Conclusion

In conclusion, automatic analyses of MRI scans of MS patients may provide more accurate and faster assessments than the traditional evaluation performed by the neuro-radiologist. LQ seems like a promising supplement to the evaluation by the neuro-radiologist, providing an automated tool for assessment of MS lesions and brain volume in MS patients.

## 6 Ethics statement

This study was carried out in accordance with the recommendations of the Regional Committee for Medical and Health Research Ethics with written informed consent from all subjects. All subjects gave written informed consent in accordance with the Declaration of Helsinki. The South Eastern Regional Committee for Medical and Health Research Ethics approved the protocol.

## Data Availability

Due to the sensitive nature of the data, they can not be freely downloaded from a website. Please contact the Authors if you wish to ask for Access to these data, or want to collaborate With us.

## 7 Acknowledgements

We thank all the patients who participated in our study. We acknowledge the collaboration with members of the Multiple Sclerosis Research Group at the University of Oslo and Oslo University hospital, especially Professor Elisabeth G. Celius. We acknowledge the collaboration with the Regional Core Facility in Translational MRI with leader Frode A. Tuvnes, Lisa Kjønigsen and research assistant Eva B. Aamodt.

## 8 Author contributions statement

SB, EH, PBH, HFH and MKB contributed to the conception and design of the study. SB, EAH, VS, GON, PBH, HFH, PS and MKB contributed to the acquisition and analysis of data. SB, EAH, PBH, HFH and MKB drafted the text and figures. During review and editing of this manuscript, all authors contributed.

## 10 Contribution to the field

Visual examination of MRI scans is currently the standard procedure for the diagnosis and follow-up of MS patients. In this paper we show that using a program to automatically detect brain white matter lesions in patients with MS, has a high correlation with the results of counting the same lesions performed by a neuro-radiologist. The LQ program in addition provides lesion volumes and several volumes of brain grey matter, which is not part of a visual evaluation of brain MRI scans. This additional information may prove to be important for future correlation with clinical status and prognosis of the patients. We speculate that using an automated tool like this, in combination with an evaluation by a neuro-radiologist, will be the future standard procedure in the follow-up of MS. In order to start using these tools, we need more experience and more research on their advantages and drawbacks compared to current practice.

### 11 Data availability statement

The current dataset cannot be made publicly available for ethical reasons, and public availability would compromise patient confidentiality and participant privacy. The study was conducted in humans and the dataset includes sensitive and personal information on individuals. A portion of data can be made available upon request to interested, qualified researchers provided that an agreement is made up. The minimal data set will enable replication of the reported study findings. Requests to access the datasets should be directed to professor Hanne F Harbo.

